# Kidney Function Variability and Cardiovascular Outcomes in Heart Failure: Effects of Empagliflozin in the EMPEROR-Reduced Trial

**DOI:** 10.64898/2026.11.16.26368172

**Authors:** Masayuki Kunitake, Kyoko Higuchi, Yoshiro Sakai, Yoshihiro Fukumoto

## Abstract

**Background:** Although sodium-glucose cotransporter-2 (SGLT2) inhibitors improve cardiovascular and kidney outcomes in patients with heart failure (HF), the prognostic significance of intraindividual kidney function variability and its modification by SGLT2 inhibition remain unclear. We investigated these relationships in patients with HF with reduced ejection fraction (HFrEF).

**Methods:** This secondary analysis included 3,492 patients from the EMPEROR-Reduced trial. A location-scale joint model (LSJM) simultaneously evaluated the associations of estimated glomerular filtration rate (eGFR) levels, longitudinal slopes, and intraindividual variability from day 28 onward with the composite outcome of cardiovascular death or HF hospitalization. Patients were stratified into tertiles of estimated eGFR variability using common cutoffs across the empagliflozin and placebo groups, allowing comparisons of cardiovascular outcomes at comparable variability levels.

**Results:** Greater intraindividual eGFR variability was independently associated with increased cardiovascular risk (hazard ratio: 1.08 per 1 mL/min/1.73 m² increase in estimated standard deviation; P < 0.001). The model incorporating eGFR levels, slopes, and variability demonstrated the best fit. Empagliflozin significantly reduced eGFR variability compared with placebo (standard deviation ratio: 0.93; P = 0.005) and attenuated annual eGFR decline by 1.83 mL/min/1.73 m²/year, despite an initial eGFR reduction of approximately 2.5 mL/min/1.73 m². Event-free survival progressively decreased with increasing eGFR variability in both treatment groups. Importantly, within each variability tertile defined by common cutoffs, empagliflozin-treated patients consistently exhibited higher event-free survival than placebo-treated patients.

**Conclusion:** In patients with HFrEF, greater intraindividual eGFR variability was independently associated with adverse cardiovascular outcomes, beyond eGFR levels and slopes. Empagliflozin reduced kidney function variability and attenuated long-term eGFR decline, with more favorable cardiovascular outcomes observed even among patients with comparable levels of kidney function variability.

## Introduction

Chronic heart failure (HF) is a leading cause of hospitalization, particularly among older adults.^1–3^ Traditionally, HF has been classified into three categories based on left ventricular ejection fraction (LVEF): HF with reduced (HFrEF; LVEF <40%), mildly reduced (HFmrEF; LVEF 40–49%), and preserved ejection fraction (HFpEF; LVEF ≥50%).^4–6^ However, the updated 2026 European Society of Cardiology (ESC) guidelines have proposed a simplified classification into HFrEF (LVEF <50%) and HFpEF (LVEF ≥50%), based on LVEF and clinical evidence of HF.^7^

HF and chronic kidney disease (CKD) frequently coexist because of shared risk factors, including diabetes mellitus, hypertension, and atherosclerotic cardiovascular disease, as well as altered renal hemodynamics, sodium and water retention, and neurohormonal dysregulation. These conditions exhibit complex bidirectional pathophysiological and clinical interactions.^8^ Consequently, the assessment and management of kidney function are essential components of comprehensive HF care.

Large randomized clinical trials, including EMPEROR-Reduced and DAPA-HF, have demonstrated that sodium-glucose cotransporter-2 (SGLT2) inhibitors reduce the risk of cardiovascular events and attenuate the long-term decline in estimated glomerular filtration rate (eGFR) in patients with HF.^9–14^ However, initiation of SGLT2 inhibitor therapy is frequently accompanied by a transient reduction in eGFR, commonly referred to as the “initial dip,” which is primarily attributed to hemodynamic changes that reduce intraglomerular pressure.^13,15–18^ Although this early decline is generally considered a physiological response associated with subsequent kidney protection, concerns remain regarding its potential relationship with progressive kidney dysfunction in certain patients.^18^ Furthermore, the prognostic significance of the magnitude of the initial dip and the subsequent level of kidney function remains incompletely understood.

Previous studies have reported that fluctuations in kidney function are associated with adverse clinical outcomes, including cardiovascular mortality, in patients with HF.^19^ In particular, intraindividual eGFR variability during the chronic maintenance phase may reflect underlying hemodynamic instability, including reduced cardiac output and renal congestion.^20–22^ Nevertheless, the effects of SGLT2 inhibitors on such kidney function instability have not been adequately investigated.

Serum creatinine levels and eGFR are inherently subject to physiological fluctuations related to changes in volume status, dietary intake, and intercurrent illness, as well as measurement variability. Consequently, distinguishing transient, reversible hemodynamic changes from progressive, irreversible kidney injury based on measurements obtained at a single time point is challenging. Importantly, the prognostic significance of intraindividual eGFR variability following SGLT2 inhibitor initiation remains unclear.

The present study aimed to determine whether kidney function stability, assessed by longitudinal eGFR variability, is associated with the risk of cardiovascular death or hospitalization for HF in patients with HFrEF, and to investigate how empagliflozin influences kidney function stability and cardiovascular outcomes. To address these objectives, we applied a location-scale joint model (LSJM)^23^ to individual participant data from the EMPEROR-Reduced trial, integrating longitudinal eGFR measurements with cardiovascular outcomes.

## Methods

### Study Design and Data Source

This study was a secondary analysis of individual participant data from the EMPEROR-Reduced trial, provided by Boehringer Ingelheim through the Vivli data-sharing platform.^10^ The EMPEROR-Reduced trial was a randomized, double-blind, placebo-controlled trial involving 3,730 patients with HFrEF. Participants were randomly assigned to receive empagliflozin or placebo, with stratification according to geographical region, diabetes status at screening, and baseline eGFR (≥60 vs. <60 mL/min/1.73 m²). In the original trial, empagliflozin significantly reduced the risk of the primary composite outcome of cardiovascular death or hospitalization for HF compared with placebo (hazard ratio [HR], 0.75; 95% confidence interval [CI], 0.65–0.86).

### Study Population

The present analysis included patients enrolled in the EMPEROR-Reduced trial who received at least one dose of empagliflozin or placebo. Patients were excluded if information on the randomization stratification factors was missing, if eGFR measurements at baseline or day 28 were unavailable, or if follow-up data for cardiovascular outcomes were incomplete.

### Cardiovascular Outcomes

The primary outcome was the composite endpoint defined in the original EMPEROR-Reduced trial: time from treatment initiation to the first occurrence of cardiovascular death or hospitalization for worsening heart failure, as determined according to the trial’s endpoint adjudication procedures. Patients who did not experience a primary outcome event during follow-up, were lost to follow-up, or died from non-cardiovascular causes were right-censored at the date of their last available follow-up or death, as appropriate. For the landmark analyses, the time origin was set at day 28 after treatment initiation.

### Statistical analysis

Baseline characteristics were summarized as medians with interquartile ranges (IQRs) for continuous variables and as frequencies and percentages for categorical variables.

To address the study objectives, we evaluated three interrelated aspects of kidney function. First, we assessed the prognostic significance of eGFR levels at day 28, following the initial treatment-associated decline in eGFR. Second, we investigated whether intraindividual eGFR variability during the maintenance phase was associated with subsequent cardiovascular events, independently of eGFR levels and longitudinal slopes. Third, we evaluated the effects of empagliflozin on eGFR levels, slopes, and intraindividual variability, and examined the associations of these kidney function characteristics with cardiovascular outcomes.

To comprehensively evaluate these relationships, we applied a LSJM, which integrates longitudinal eGFR trajectories, intraindividual variability, and time-to-event outcomes while accounting for the covariance between random effects in the longitudinal mean (location) and variability (scale) components.

To distinguish the acute hemodynamic reduction in eGFR following SGLT2 inhibitor initiation (the initial dip) from subsequent chronic changes in kidney function, we performed a landmark analysis using day 28 after treatment initiation as the time origin (time = 0). A linear mixed-effects model was constructed for the longitudinal eGFR trajectory, incorporating time since day 28 (in years), treatment assignment (empagliflozin vs. placebo), and the time-by-treatment interaction as fixed effects, together with patient-specific random intercepts and random slopes.

To account for heterogeneity in within-patient eGFR variability and its potential changes over time, we simultaneously specified a scale submodel for the residual standard deviation of repeated eGFR measurements. This submodel included time and treatment assignment as fixed effects and a patient-specific random intercept to capture between-patient heterogeneity in intraindividual variability.

The longitudinal and survival submodels were jointly linked through three time-dependent characteristics of kidney function:

1. Current value (location): The estimated absolute eGFR level at each time point.
2. Current slope (derivative of location): The estimated instantaneous rate of change in eGFR at each time point.
3. Current variability (scale): The estimated magnitude of intraindividual eGFR variability, expressed as the residual standard deviation at each time point.

These characteristics were simultaneously incorporated into the survival submodel through association functions, allowing us to quantify the association between eGFR variability and subsequent cardiovascular events independently of the current eGFR level and slope.

To evaluate model fit, we constructed and compared three models: a current-value model (CV model), incorporating only the current eGFR value; a current-value-and-slope model (CVS model), incorporating both the current eGFR value and slope; and a current-value-slope-and-variability model (CVSV model), incorporating all three longitudinal characteristics. Model fit was compared using the Akaike information criterion (AIC).

For the survival submodel, the baseline hazard function was modeled using a B-spline function with one internal knot. The model was adjusted for treatment assignment (empagliflozin vs. placebo) and the randomization stratification factors, including baseline eGFR category (≥60 vs. <60 mL/min/1.73 m²), diabetes status, and geographical region. The time origin for the survival submodel was also set at day 28.

As a sensitivity analysis, we assessed whether the findings from the primary analysis were robust to the selection of baseline covariates in the survival submodel. Specifically, we refitted the CVSV model for the primary composite outcome after excluding baseline covariates, including geographical region, prediabetes, type 2 diabetes, and baseline eGFR category, retaining treatment assignment as the only covariate in the survival submodel. The specifications of the longitudinal location submodel, scale submodel, shared random-effects structure, and baseline hazard function were identical to those used in the primary analysis.

Longitudinal eGFR measurements obtained on days 28, 85, 225, 365, 533, and 701 were used to characterize kidney function trajectories. Model-estimated longitudinal eGFR trajectories were graphically displayed (**Figure 1**). To assess the agreement between model-based hazard predictions and observed event rates, we plotted the mean and median of individual predicted cumulative hazard curves alongside the cumulative hazard estimated for the overall study population using the Nelson–Aalen estimator (**Figure 2**).

**Figure 1.** Observed and Predicted Estimated Glomerular Filtration Rate by Treatment Groups. Points and error bars represent observed mean eGFR and standard deviations (SD) centered at Day 28 post-drug initiation. Solid lines represent population-level predicted eGFR trajectories derived from the Location-Scale Joint Model (CVSV model). Blue represents the Placebo group, and red represents the Empagliflozin group. ***Abbreviations****: eGFR, estimated glomerular filtration rate; SD, standard deviation*.

**Figure 2.** Model-Predicted Cumulative Hazard Curves Compared with Nelson–Aalen Estimates. Individual predicted cumulative hazard trajectories for all subjects are shown in gray lines. The red solid line and blue dashed line represent the mean and median predicted cumulative hazards, respectively. The black solid line represents the Nelson–Aalen estimates (observed data), demonstrating strong agreement between model predictions and observed survival data.

Furthermore, using the fitted LSJM, we estimated the patient-specific standard deviation of eGFR variability at day 28. All patients, irrespective of treatment assignment, were stratified into tertiles (low, intermediate, and high) based on the distribution of estimated eGFR variability in the overall study population, using common tertile cutoffs for both treatment groups. Individual longitudinal eGFR trajectories were then visualized using spaghetti plots, and Kaplan–Meier survival curves were constructed for the six subgroups defined by treatment assignment (empagliflozin or placebo) and eGFR variability tertile.

All statistical analyses were performed using R version 4.4.0 (R Core Team, 2024) and RStudio version 2024.04.2 (Posit Software, PBC). The LSJM analyses were conducted using the R package *FlexVarJM*.

## Results

### Study Population and Baseline Characteristics

Of the 7,220 patients screened for the EMPEROR-Reduced trial, 3,730 were randomized. After excluding patients with missing information on randomization stratification factors, those who did not receive empagliflozin or placebo, those without eGFR measurements at baseline or day 28, and those with incomplete follow-up data for cardiovascular outcomes, 3,492 patients were included in the present analysis. Baseline characteristics of the study population are summarized in **Table 1**. Patient characteristics were well balanced between the empagliflozin and placebo groups.

**Table 1.** Baseline Demographics and Clinical Characteristics of the Study Population.

| Characteristic | **Overall**<br>**N = 3,492<sup>1</sup>** | **Placebo**<br>**N = 1,731<sup>1</sup>** | **Empagliflozin**<br>**N = 1,761<sup>1</sup>** |
|---|---|---|---|
| **Age (years)** | 68 (60, 75) | 68 (60, 75) | 68 (61, 75) |
| Unknown | 5 | 1 | 4 |
| **Sex** |  |  |  |
| Feale | 828 (24%) | 415 (24%) | 413 (23%) |
| Male | 2,664 (76%) | 1,316 (76%) | 1,348 (77%) |
| **Geographical Region** |  |  |  |
| Asia | 469 (13%) | 229 (13%) | 240 (14%) |
| Europe | 1,275 (37%) | 635 (37%) | 640 (36%) |
| Latin America | 1,195 (34%) | 591 (34%) | 604 (34%) |
| North America | 389 (11%) | 191 (11%) | 198 (11%) |
| Other | 164 (4.7%) | 85 (4.9%) | 79 (4.5%) |
| **Body Mass Index (kg/m<sup>2</sup>)** | 27.3 (24.1, 31.1) | 27.2 (24.0, 30.9) | 27.4 (24.2, 31.2) |
| **Alcohol Consumption** |  |  |  |
| Does not drink alcohol | 2,238 (64%) | 1,091 (63%) | 1,147 (65%) |
| Drinks alcohol (no interference with participation) | 1,112 (32%) | 568 (33%) | 544 (31%) |
| Unknown | 142 (4.1%) | 72 (4.2%) | 70 (4.0%) |
| **Smoking Status** |  |  |  |
| Currently smoker | 355 (10%) | 171 (9.9%) | 184 (10%) |
| Ex-smoker | 1,560 (45%) | 768 (44%) | 792 (45%) |
| Never smoked | 1,455 (42%) | 734 (42%) | 721 (41%) |
| Unknown | 122 (3.5%) | 58 (3.4%) | 64 (3.6%) |
| **Diabetes Status** |  |  |  |
| Normal (no pre-diabetes) | 575 (16%) | 287 (17%) | 288 (16%) |
| Pre-diabetes | 1,184 (34%) | 582 (34%) | 602 (34%) |
| Type 2 Diabetes | 1,733 (50%) | 862 (50%) | 871 (49%) |
| **Hypertension** | 2,527 (72%) | 1,245 (72%) | 1,282 (73%) |
| **NYHA Functional Class** |  |  |  |
| II | 2,649 (76%) | 1,312 (76%) | 1,337 (76%) |
| III | 826 (24%) | 410 (24%) | 416 (24%) |
| IV | 17 (0.5%) | 9 (0.5%) | 8 (0.5%) |
| **Left Ventricular Ejection Fraction (%)** | 28 (24, 31) | 28 (24, 31) | 28 (25, 32) |
| **Hemoglobin A1c (%)** | 6.10 (5.70, 6.90) | 6.10 (5.70, 6.90) | 6.10 (5.70, 6.90) |
| Creatinine_Clearance_bl | 60 (45, 78) | 60 (45, 78) | 60 (44, 78) |
| Renin-angiotensin system inhibitor | 3,068 (88%) | 1,519 (88%) | 1,549 (88%) |

Data are presented as Median (Q1, Q3) for continuous variables and n (%) for categorical variables.
**Abbreviations:** NYHA, New York Heart Association.

### Model Fit and Longitudinal eGFR Trajectories

**Table 2** presents the parameter estimates from the CV, CVS, and CVSV models. Estimates for the mean structure of the longitudinal submodel and the eGFR standard deviation ratio, representing intraindividual variability, were largely consistent across the three models. Comparison of model fit using the AIC demonstrated that the CVSV model had the lowest AIC, indicating the best relative fit to the observed data. The regression coefficients and 95% confidence intervals estimated using the CVSV model remained largely unchanged in the sensitivity analysis excluding baseline covariates from the survival submodel, supporting the robustness of the findings (**Table S1**).

**Table 2.**
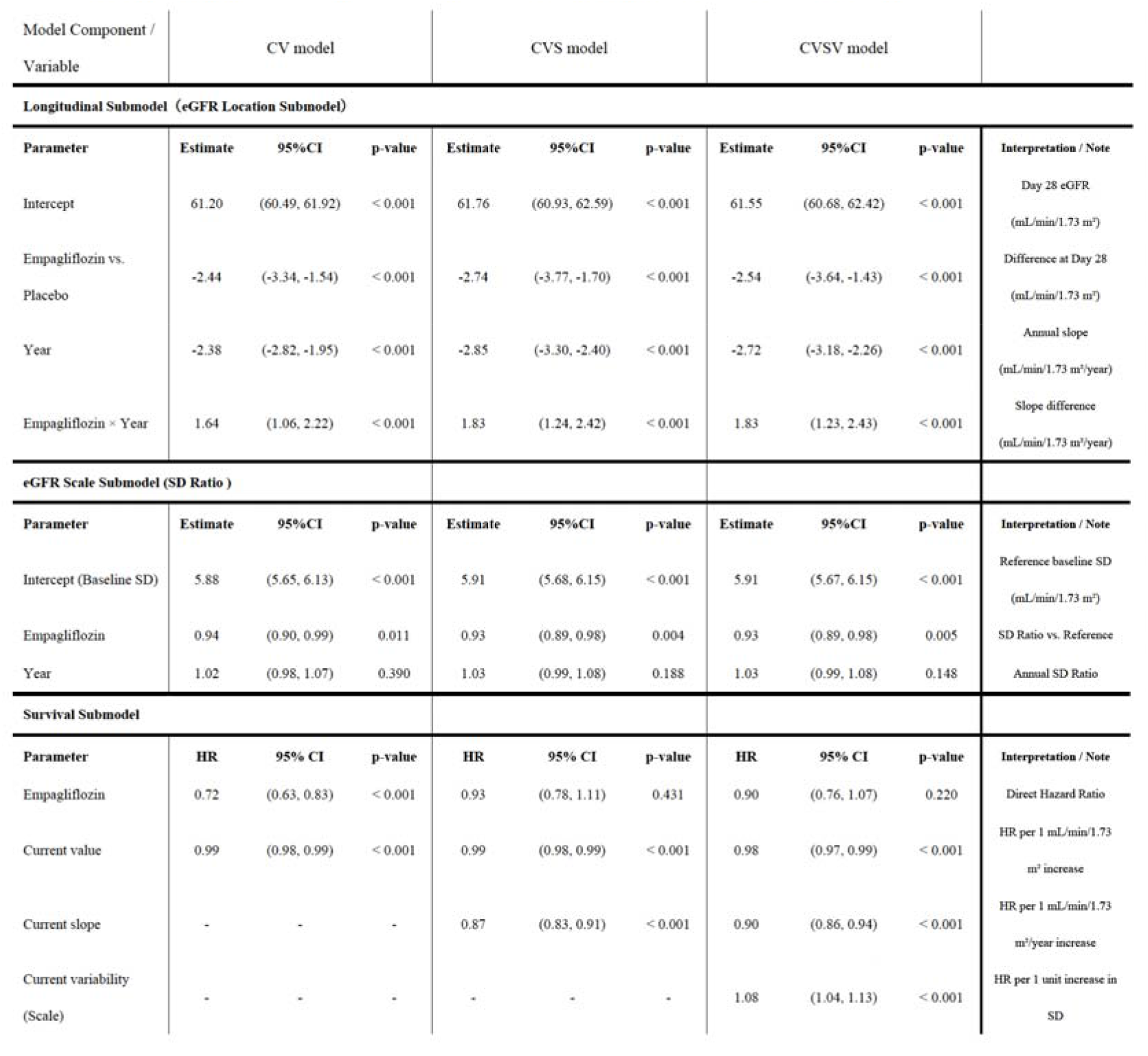
Parameter Estimates from Location-Scale Joint Models (LSJM) Centered at Day 28 Post-Drug Initation.

In the best-fitting CVSV model, the estimated eGFR at day 28 was approximately 2.5 mL/min/1.73 m² lower in the empagliflozin group than in the placebo group, consistent with the early treatment-associated decline in eGFR (initial dip). However, the subsequent annual rate of eGFR decline was −2.72 mL/min/1.73 m²/year in the placebo group, whereas the decline was attenuated by 1.83 mL/min/1.73 m²/year in the empagliflozin group. Consequently, the estimated eGFR trajectories crossed approximately 1.5 years after treatment initiation, with higher eGFR values thereafter in the empagliflozin group (**Table 2** and **Figure 1**). Furthermore, intraindividual eGFR variability was significantly lower in the empagliflozin group than in the placebo group, indicating greater stability of kidney function during empagliflozin treatment.

### Associations With Cardiovascular Events and Model Validation

In the survival submodel, lower absolute eGFR levels, steeper eGFR declines, and greater intraindividual eGFR variability were each independently and significantly associated with an increased hazard of the composite cardiovascular outcome. After accounting for these kidney function characteristics, empagliflozin treatment remained associated with a numerically lower hazard of cardiovascular events; however, this association was not statistically significant (P = 0.22).

**Figure 2** shows the mean and median of the individual predicted cumulative hazard curves, together with the population-level cumulative hazard curve estimated using the Nelson–Aalen method. The mean and median cumulative hazard curves predicted by the CVSV model closely approximated the nonparametric Nelson–Aalen estimate, suggesting good agreement between model-based predictions and observed cumulative hazards.

### Cardiovascular Outcomes According to eGFR Variability

Using the fitted CVSV model, all patients, irrespective of treatment assignment, were stratified into tertiles of estimated intraindividual eGFR variability (low, intermediate, and high), based on the patient-specific residual standard deviation at day 28. The same tertile cutoffs were applied to both treatment groups, allowing comparisons between empagliflozin- and placebo-treated patients with comparable levels of eGFR variability. Individual eGFR trajectories were then visualized separately for the empagliflozin and placebo groups within each variability tertile (**Figure 3**). Patients in higher variability tertiles exhibited greater fluctuations in their longitudinal eGFR trajectories, reflecting less stable kidney function over time.

**Figure 3.** Observed eGFR Trajectories by Treatment Arm and Model-Predicted Variability Tertile. Individual longitudinal trajectories of estimated glomerular filtration rate (eGFR) from day 28 onward, stratified by treatment assignment (empagliflozin or placebo) and tertiles of model-predicted intraindividual eGFR variability. Variability was estimated as the patient-specific residual standard deviation at day 28 using the location-scale joint model (LSJM). Tertiles were defined using common cutoffs across the overall study population (low: <4.00; intermediate: 4.00–5.53; high: ≥5.53 mL/min/1.73 m²), allowing comparisons between treatment groups with comparable levels of estimated eGFR variability. Each colored line represents an individual patient’s observed eGFR trajectory. Greater fluctuations in eGFR were observed in higher variability tertiles in both treatment groups.

Kaplan–Meier analyses were subsequently performed to evaluate event-free survival across six groups defined by treatment assignment and tertiles of estimated eGFR variability (**Figure 4**). In both treatment groups, event-free survival progressively decreased with increasing eGFR variability, from the low to intermediate and high tertiles. The placebo group with high eGFR variability exhibited the poorest prognosis, with the most rapid decline in event-free survival. Within each variability tertile, patients receiving empagliflozin consistently demonstrated higher event-free survival probabilities than those receiving placebo.

**Figure 4.** Kaplan-Meier Curves for Cardiovascular Events by Treatment Arm and Model-Predicted eGFR Variability Tertile. Kaplan–Meier curves showing event-free survival for the composite outcome of cardiovascular death or hospitalization for heart failure, stratified by treatment assignment (empagliflozin or placebo) and tertiles of model-predicted intraindividual estimated glomerular filtration rate (eGFR) variability. Variability was estimated as the patient-specific residual standard deviation at day 28 using the location-scale joint model (LSJM). Tertiles were defined using common cutoffs across the overall study population (low: <4.00; intermediate: 4.00–5.53; high: ≥5.53 mL/min/1.73 m²), allowing comparisons between treatment groups with comparable levels of estimated eGFR variability. Event-free survival progressively decreased with increasing eGFR variability in both treatment groups. Within each variability tertile, empagliflozin-treated patients exhibited higher event-free survival probabilities than placebo-treated patients. The numbers of patients at risk are shown below the curves.

## Discussion

In this secondary analysis of individual participant data from the EMPEROR-Reduced trial, we demonstrated that greater intraindividual variability in kidney function was associated with an increased risk of cardiovascular death or hospitalization for heart failure in patients with HFrEF. Furthermore, empagliflozin treatment was associated with lower eGFR variability and more favorable cardiovascular outcomes across different levels of estimated kidney function variability. By applying a LSJM to longitudinal eGFR measurements and cardiovascular outcomes from day 28 onward, we comprehensively evaluated three dimensions of kidney function: absolute eGFR levels, longitudinal slopes, and intraindividual variability.

The CVSV model demonstrated the lowest AIC compared with models incorporating the current eGFR value alone (CV model) or the current value and slope (CVS model), indicating improved model fit when intraindividual variability was included. These findings suggest that kidney function stability provides additional prognostic information beyond conventional measures of kidney function in patients with HFrEF.

Notably, after accounting for eGFR levels, slopes, and intraindividual variability, the association between empagliflozin treatment and cardiovascular outcomes was attenuated and no longer statistically significant (HR, 0.90; 95% CI, 0.76–1.07; P = 0.220). Although this finding raises the possibility that favorable changes in kidney function may contribute to the cardiovascular benefits of empagliflozin, the present analysis does not establish a causal mediating effect.

### Prognostic Significance of Intraindividual eGFR Variability

Previous studies have demonstrated that worsening kidney function during HF treatment is associated with adverse clinical outcomes.^24^ However, the prognostic significance of intraindividual eGFR variability during the maintenance phase of HF treatment remains incompletely understood. Increased visit-to-visit eGFR variability in patients with HF or chronic kidney disease (CKD) has been proposed as a potential marker of hemodynamic instability, reflecting impaired glomerular autoregulation, renal microvascular dysfunction, or fluctuations in renal perfusion associated with changes in volume status.^20,21^

An important methodological strength of the present study was the application of LSJM to estimate patient-specific intraindividual eGFR variability while accounting for longitudinal eGFR trajectories and their associations with cardiovascular outcomes. When patients were stratified into tertiles according to their model-estimated eGFR standard deviation at day 28, individual eGFR trajectories demonstrated progressively greater fluctuations from the low- to high-variability tertiles (**Figure 3**). Furthermore, the CVSV model demonstrated the best relative fit among the three models evaluated (**Table 2**), supporting the additional prognostic information provided by intraindividual variability.

Importantly, greater estimated eGFR variability was independently associated with an increased hazard of cardiovascular events in the survival submodel (HR, 1.08 per 1 mL/min/1.73 m² increase in estimated residual standard deviation; P < 0.001). Consistent with this finding, Kaplan–Meier analyses demonstrated a stepwise reduction in event-free survival across increasing tertiles of eGFR variability (**Figure 4**). These findings highlight kidney function instability as a potentially important prognostic marker, beyond absolute eGFR values and the rate of kidney function decline.

### Effects of Empagliflozin on Kidney Function Stability and Cardiovascular Outcomes

In the present CVSV model, empagliflozin significantly reduced intraindividual eGFR variability compared with placebo (standard deviation ratio, 0.93; P = 0.005), corresponding to a 7% reduction in the estimated residual standard deviation. This finding suggests that empagliflozin may not only attenuate long-term kidney function decline but also promote greater stability of kidney function during maintenance therapy.

Potential mechanisms include improved regulation of intraglomerular pressure, more stable volume status, and reduced fluctuations in renal hemodynamics. Although these mechanisms were not directly assessed in the present study, they provide plausible physiological explanations for the observed reduction in eGFR variability.

Furthermore, when patients were stratified into tertiles of estimated eGFR variability using common cutoffs across both treatment groups, empagliflozin-treated patients consistently exhibited more favorable event-free survival than placebo-treated patients within each variability tertile (**Figure 4**). Thus, even among patients with comparable levels of estimated kidney function variability, empagliflozin treatment was associated with better cardiovascular outcomes.

Based on the estimated association between eGFR variability and cardiovascular risk, the reduction in variability observed with empagliflozin could contribute to a modest decrease in cardiovascular event hazard, independently of absolute eGFR levels and longitudinal slopes. However, these findings do not establish that the cardiovascular benefits of empagliflozin are causally mediated by reduced eGFR variability. Further studies are warranted to clarify whether improved kidney function stability represents a mechanism contributing to cardiovascular protection.

### Clinical Implications of the Initial eGFR Dip and Long-Term Improvement in eGFR Slope

Our findings also provide clinically relevant insights into the relationship between the initial eGFR dip following empagliflozin initiation and subsequent long-term changes in kidney function. In the longitudinal submodel, treatment assignment was incorporated into the eGFR intercept at day 28, whereas time and the time-by-treatment interaction were used to estimate subsequent eGFR slopes. Given the randomized design of the original trial and the overall balance in baseline kidney function between treatment groups, the estimated between-group difference in eGFR at day 28 may largely reflect the early treatment-associated decline in eGFR.

In the present analysis, empagliflozin was associated with an approximately 2.54 mL/min/1.73 m² lower eGFR at day 28 than placebo. Based on the estimated association between the current eGFR value and cardiovascular risk, this difference corresponded to an approximately 5% higher modeled hazard. In contrast, empagliflozin attenuated the subsequent annual eGFR decline by 1.83 mL/min/1.73 m²/year compared with placebo. The model-based association for a corresponding difference in eGFR slope was equivalent to an approximately 17% lower hazard. These estimates suggest that the relatively modest early reduction in eGFR associated with empagliflozin initiation should be considered in the context of its favorable long-term kidney function trajectory.

Nevertheless, a substantially greater early decline in eGFR may warrant closer clinical attention. For example, a hypothetical 10 mL/min/1.73 m² lower eGFR, evaluated using the estimated current-value association, would correspond to an approximately 22% higher modeled hazard. However, this calculation does not establish that an initial eGFR dip of this magnitude directly causes an increase in cardiovascular risk, nor does it identify a validated threshold for treatment discontinuation. In patients experiencing a pronounced early decline in eGFR, careful assessment of volume status, blood pressure, concomitant medications, and other potential causes of kidney dysfunction remains important.

Overall, these findings support interpreting early changes in eGFR within the broader context of longitudinal kidney function trajectories rather than relying on a single measurement. A modest initial eGFR dip should not necessarily be interpreted as progressive kidney injury, particularly when subsequent kidney function remains stable.

### Limitations

Several limitations should be acknowledged. First, the present analysis was restricted to patients with HFrEF enrolled in the EMPEROR-Reduced trial. Because cardiorenal hemodynamics and underlying risk profiles may differ in patients with HFpEF or CKD without HF, caution is warranted when generalizing these findings to other populations. Second, the analysis used day 28 after treatment initiation as the landmark time point. Consequently, patients who died, experienced an early primary outcome event, or discontinued treatment before day 28 and were therefore ineligible for the analysis were not represented in the landmark cohort. In addition, the initial change in eGFR from baseline to day 28 was assessed using only two measurements, preventing a detailed characterization of the timing, magnitude, and recovery of the early eGFR dip. Third, longitudinal eGFR measurements were obtained at prespecified study visits rather than through continuous monitoring. Therefore, transient changes in kidney function occurring between visits, particularly during episodes of acute clinical deterioration, may not have been fully captured. Changes in concomitant medications, including diuretics and renin–angiotensin system inhibitors, as well as informative missingness or dropout, may also have influenced the estimated eGFR slopes and intraindividual variability. Furthermore, although the LSJM accounted for longitudinal eGFR trajectories and their variability, concurrent changes in other clinical parameters, such as blood pressure, body weight, and volume status, and their potential interactions with kidney function were not fully incorporated into the model. Finally, eGFR was estimated from serum creatinine concentrations, which are influenced by nonrenal factors, including muscle mass, dietary intake, and hydration status. Accordingly, some of the estimated intraindividual variability may reflect physiological or measurement-related fluctuations rather than changes in intrinsic kidney function. Future studies incorporating complementary markers, such as cystatin C, and external validation in independent HF and CKD cohorts are warranted.

## Conclusions

In patients with HFrEF, greater intraindividual eGFR variability was independently associated with an increased risk of cardiovascular death or HF hospitalization, beyond absolute eGFR levels and longitudinal slopes. Empagliflozin significantly reduced eGFR variability and attenuated long-term eGFR decline despite an initial eGFR dip. These findings highlight kidney function stability as a potentially important prognostic marker and provide further insight into the cardiovascular and kidney benefits of SGLT2 inhibition.

## Data Availability

All data produced are available online at Vivli.

## IRB information

The requirement for ethical approval was waived by the Ethical Committee of Kurume University because this study used previously collected, de-identified clinical trial data.

## Abbreviations list

AIC: Akaike information criterion
CI: confidence interval
CKD: chronic kidney disease
CV: current-value
CVS: current-value-and-slope
CVSV: current-value-slope-and-variability
eGFR: estimated glomerular filtration rate
ESC: European Society of Cardiology
HF: heart failure
HFmrEF: HF with mildly reduced ejection fraction
HFpEF: HF with preserved ejection fraction
HFrEF: HF with reduced ejection fraction
HR: hazard ratio
IQRs: interquartile ranges
LSJM: location-scale joint model
LVEF: left ventricular ejection fraction
SGLT2: sodium-glucose cotransporter 2.

## Acknowledgments

This study is based on research using data from data contributors Boehringer Ingelheim Co., Ltd. that has been made available through Vivli, Inc. Vivli has not contributed to or approved, and is not in any way responsible for, the contents of this publication.

## Data availability statement

The data underlying this article will be shared with others for the purpose of reproducing results or replicating procedures upon reasonable request to the corresponding author, subject to institutional and ethics committee approval.

## Authors’ Contributions

Masayuki Kunitake: Conceptualization, Methodology, Formal analysis, Data Curation, and Writing-Original draft preparation.

Kyoko Higuchi: Methodology, Data Curation, Formal analysis, Investigation, Writing-Review and editing..

Yoshiro Sakai: Methodology, Investigation.

Yoshihiro Fukumoto: Project administration, Conceptualization, Writing-Review and editing.

All authors have read and approved the final article.

